# Identifying Opioid-Related Pharmacogenomic Variants in High-Risk Hospitalized Infants: A Pilot Study

**DOI:** 10.1101/2024.10.25.24316140

**Authors:** Rabab M. Barq, Shadassa Ourshalimian, Simran Maggo, Olivia A. Keane, Jenny Q. Nguyen, Matthew A. Deardorff, Tamorah Lewis, Ashwini Lakshmanan, Scott A. Mosley, Lorraine I. Kelley-Quon

## Abstract

**Introduction:** Pharmacogenomic (PGx) variants impact the pharmacodynamics and pharmacokinetics of opioids and the brain’s reward, cognition, stress, and pain pathways. This study examines the prevalence of PGx variants impacting opioid response among a cohort of high-risk infants.

**Methods:** This retrospective study was conducted at a quaternary children’s hospital from 2009-2020. Infants <1y with ≥1 high-risk condition (congenital heart disease (CHD), medical or surgical necrotizing enterocolitis (NEC), thoracoabdominal surgery, very or extremely low birthweight, hypoxic ischemic encephalopathy, or extracorporeal membrane oxygenation) with exome sequencing were included. Prevalence of opioid-related variants (*COMT, DRD2/ANKK1, ABCB1, OPRM1, CYP2B6, CYP2D6*) were compared to Ensembl, a database of published genomic cohorts.

**Results:** Overall, 111 high-risk infants were identified (62.2% male, 47.7% Hispanic/Latino, 18.0% premature, and 82.0% CHD). Most underwent surgery (68.2%), with 37.8% undergoing CHD surgery. Overall, 81.1% of infants were homozygous for ≥1 opioid-related variant(s). Compared to Ensembl, high-risk infants had a significantly higher frequency of homozygosity for *ABCB1: rs1045642* (43.6% vs. 18.7%, p<0.001) and *rs2032582* (42.7% vs. 15.9%, p<0.001). Conversely, high-risk infants had a lower frequency of homozygosity for *COMT: rs4818* (2.7% vs. 10.3%, p<0.001) and *OPRM1: rs1799971* (2.7% vs. 7.1%, p<0.001). All infants with surgical NEC (N=5) were homozygous for ≥1 opioid-related variant. The most common metabolizer phenotypes were intermediate (CYP2B6: 35.5%, CYP2D6: 20.0%) and normal (CYP2B6: 53.9%, CYP2D6: 70.9%).

**Conclusion:** Most high-risk infants carried at least one opioid-related variant, with frequencies that are significantly different from broader genetic cohorts. Larger studies inclusive of intronic PGx variants using ancestrally comparable controls are needed.

## Introduction

Pharmacogenomic (PGx) variants are known to impact the pharmacodynamics and pharmacokinetics of opioids and the brain’s reward, cognition, stress, and pain pathways [1–4]. Critically ill infants commonly receive opioids for analgesia after invasive procedures and/or sedation during mechanical ventilation [5–7]. Although opioids play an important role in pain control for hospitalized infants, higher cumulative exposures are associated with prolonged mechanical ventilation, increased total parenteral nutrition use, longer hospitalization, increased risk of methadone treatment, and costs [5–8]. Additionally, current research underscores the risk of impaired neurodevelopment associated with increasing opioid exposures during infancy [9–11].

PGx variations have been identified as a factor contributing to pain tolerance, opioid dose requirements, and risk of experiencing harmful effects of opioid medication [12–17]. Currently, there are multiple suggested PGx variants that contribute to variability in opioid metabolism, opioid response, and methadone metabolism. These include drug metabolizing enzymes (CYP2D6, CYP2B6), transporters (*ABCB1*), receptors (*OPRM1*), and central nervous system neuromodulators (*COMT, DRD2*/*ANKK1*) [2,3]. Despite the growing body of literature investigating PGx influences on opioid response, most genetic studies have been conducted in adults of European ancestry, which limits the generalizability of their findings [18]. Previous studies of pediatric patients screened for known drug response variants reveal that 97% had at least one clinically actionable PGx variant [19]. However, the prevalence of PGx variants impacting opioid response in high-risk hospitalized infants is currently unknown. The goal of this study was to describe the prevalence of opioid-related PGx variants among a diverse cohort of high-risk hospitalized infants compared to published genomic cohorts.

## Methods

### Study Design

A retrospective cohort study was conducted including high-risk infants less than one year who were admitted to a quaternary children’s hospital in an urban area between January 1, 2009, and December 31, 2020. Cohort discovery and demographic and clinical data were obtained from the Pediatric Health Information Systems (PHIS) database, an administrative database that contains inpatient data from not-for-profit, tertiary care pediatric hospitals in the United States. These hospitals are affiliated with the Children’s Hospital Association (CHA: Lenexa, KS). Data quality and reliability are assured through a joint effort between the Children’s Hospital Association and participating hospitals. CHA hospitals submit resource utilization data (e.g. pharmaceuticals, imaging, and laboratory) into PHIS. Data are de-identified at the time of data submission, and data are subjected to several reviews and validity checks before being included in the database. The study received approval from the Children’s Hospital Los Angeles Review Board (CHLA-23-00335).

To identify our cohort of high-risk infants at our institution, we searched the PHIS database using diagnostic codes from the International Classification of Diseases, Ninth Revision, Clinical Modification (ICD-9-CM) and Tenth Revision, Clinical Modification (ICD-10-CM) [8]. This cohort of infants was previously identified and published by Keane et al. 2024. Briefly, this included infants with congenital heart disease (CHD), necrotizing enterocolitis (NEC), extremely low birth weight (ELBW), very low birth weight (VLBW), hypoxemic ischemic encephalopathy (HIE), extracorporeal membrane oxygenation (ECMO), and thoracoabdominal surgery. These categories were based on the ICD-9/10 codes recorded during a patient’s encounter and are not mutually exclusive, meaning a single patient could fall into multiple categories. The Center for Personalized medicine (CPM) manages and conducts all clinical genomic sequencing at our institution, including clinical exome sequencing. High-risk infant cohort data was then merged with clinical exome sequencing data at CPM to create a comprehensive clinical and genomic dataset. Patients with ICD-9/10 codes for neonatal opioid withdrawal syndrome (NOWS) or malignancy were excluded. Infants who did not undergo a cardiac procedure and/or surgical intervention and had a congenital heart disease diagnosis code alone were excluded. Additionally, patients that received methadone alone and no other opioids and patients that stayed longer than 365 days in the hospital were excluded. Finally, patients that did not undergo clinical exome sequencing as part of their routine clinical care were excluded.

Demographic information included age at admission, sex, race, ethnicity, and insurance type. Clinical factors included the presence of prematurity (defined by the presence of a complex chronic condition flag, yes/no), use of mechanical ventilation (yes/no), surgical intervention within the first year of life (yes/no), high-risk diagnosis and/or procedure category, and the number of complex chronic conditions (CCCs; 1, 2, ≥3) for each patient [20].

### Outcome measures

Seven exonic opioid-related PGx variants previously reported in the literature were assessed including: *COMT rs:4633, rs:4680, and rs:4818* [13,21], *DRD2/ANKK1 rs1800497* [22], *ABCB1 rs1045642 and rs2032582* [23,24], *and OPRM1 rs:179971* [25] alongside CYP2B6 [26] and CYP2D6 metabolizer phenotypes [27]. Children’s Hospital Los Angeles’s Center for Personalized Medicine currently provide Clinical Laboratory Improvement Amendments and The College of American Pathologists-approved whole exome sequencing, which only covers exonic variants, therefore intronic variants were not analyzed in this study. Genotype frequencies of each genomic variant were described for our analytic sample. If an infant in our sample was homozygous for any of the seven opioid-related PGx variants, they were defined as having a PGx variant and the overall frequency of homozygosity for any variant in our analytic sample was reported. We utilized the Ensembl Genome Browser [http://www.ensembl.org/index.html] (accessed on July 3, 2024) to obtain allele frequencies and estimate prevalence of each genomic variant. Ensembl is a comprehensive genome database that includes allele frequencies for many genetic variants [28]. The prevalence of each genetic variant within our cohort was then compared to populations referenced in Ensembl. Overall prevalences combining populations of different ancestry of CYP2D6 and CYP2B6 metabolizer phenotypes are not reported in Ensembl, therefore, no comparisons between prevalences of metabolizer frequencies were made.

### Statistical Analyses

Demographic and clinical descriptive statistics were reported using counts and percentages. Chi-squared and Fischer’s exact tests were conducted when comparing categorical variables. Separate unadjusted logistic regression models were used to calculate the odds of expressing any PGx variant by high-risk category. All analyses were conducted with an α equal to 0.05. Bivariate analyses were conducted using SAS® software 9.4 (copyright © 2016 SAS Institute Inc., Cary, NC).

## Results

Following exclusions, a cohort of 111 high-risk infants who underwent clinical exome sequencing were identified (Figure 1). The cohort was 62.2% male and 47.7% was Hispanic (Table 1). Most infants were publicly insured (54.1%). Prematurity occurred in 18%, a majority underwent surgery (68.2%), and many had three or more CCCs (43.2%). CHD was present in 82% of our cohort, 37.8% underwent surgery for CHD, and 24.3% underwent non-NEC abdominal surgery. During hospitalization, 20.7% received methadone treatment.

**Table 1:** Demographic and clinical factors among high-risk infants less than one year of age.

|  | **Cohort** |  |
| --- | --- | --- |
|  | **N=111** | **%** |
| **Biologic Sex** |  |  |
| Male | 69 | 62.2 |
| Female | 42 | 37.8 |
| **Race/Ethnicity** |  |  |
| Hispanic | 53 | 47.7 |
| Non-Hispanic White | 19 | 17.1 |
| Black/African American | 10 | 9 |
| Other | 24 | 21.6 |
| Unknown | 5 | 4.5 |
| **Insurance** |  |  |
| Private | 34 | 30.6 |
| Public | 60 | 54.1 |
| Other | 17 | 15.3 |
| **Number of CCCs\*** |  |  |
| 0 | 6 | 5.4 |
| 1 | 31 | 27.9 |
| 2 | 26 | 23.4 |
| ≥3 | 48 | 43.2 |
| **High-Risk Diagnoses** |  |  |
| Congenital Heart Disease (CHD) | 91 | 82.0 |
| Surgery for CHD | 42 | 37.8 |
| Thoracoabdominal Surgery | 27 | 24.3 |
| Medical NEC | 18 | 16.2 |
| Surgical NEC | 5 | 4.5 |
| Very low birthweight | 12 | 10.8 |
| Extremely low birthweight | 7 | 6.3 |
| Hypoxic Ischemic Encephalopathy | 9 | 8.1 |
| Extracorporeal membrane oxygenation | 10 | 9.0 |
| Methadone Treatment | 23 | 20.7 |
| Homozygous for any opioid-related pharmacogenomic variant | 90 | 81.1 |
| Prematurity | 20 | 18.0 |
| Any Surgery | 69 | 62.2 |

\*CCCs = Complex Chronic Conditions

**Figure 1:**
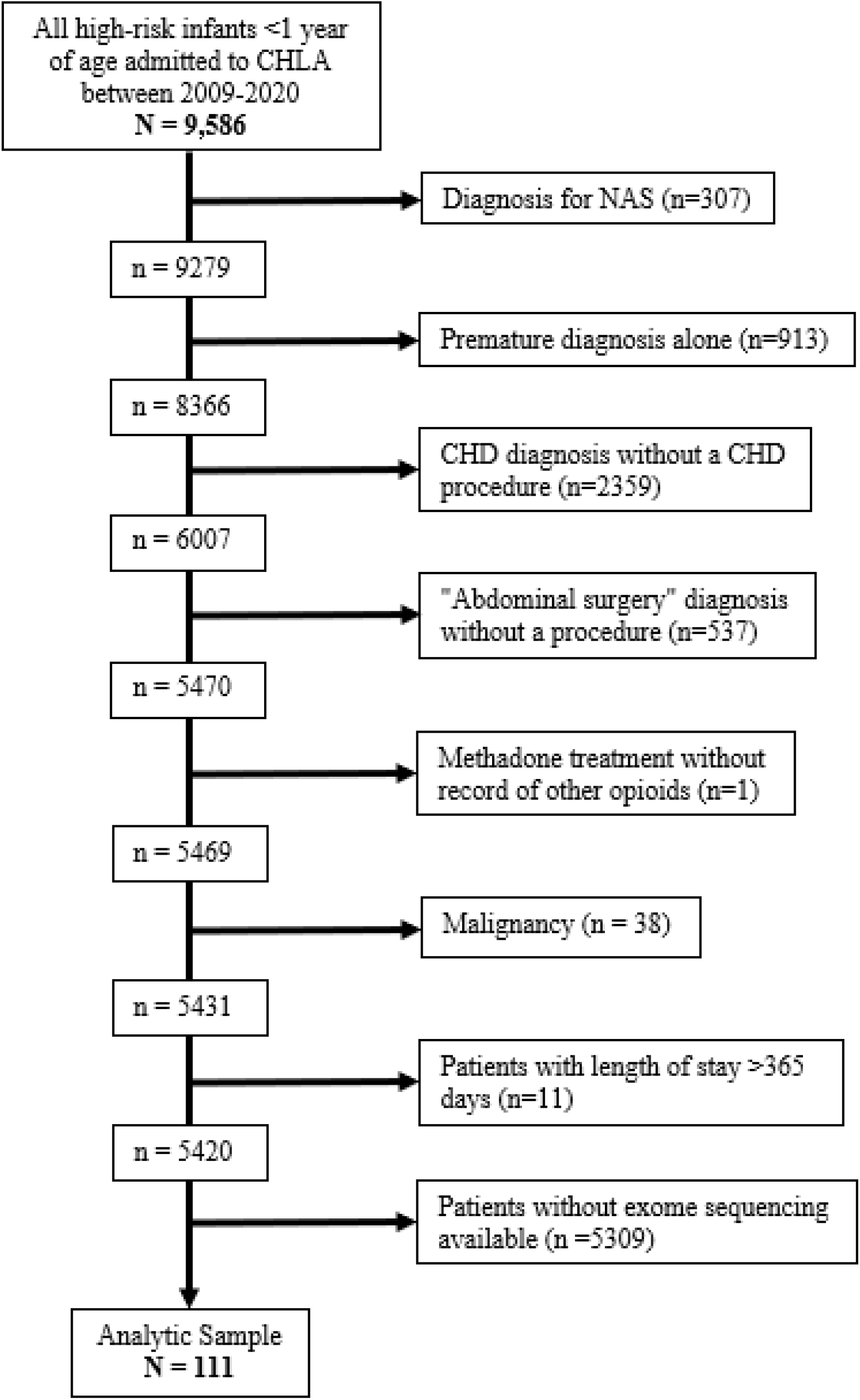
Study Flow Diagram.

Overall, 81.1% (N = 90) of infants were homozygous for at least one genomic variant associated with opioid response (Table 2). Compared to Ensembl, high-risk infants had a higher frequency of homozygosity for *ABCB1: rs1045642* (43.6% vs. 18.7%, p<0.001) and *rs2032582 (*42.7% vs. 15.9%, p<0.001). Conversely, high-risk infants had a lower frequency of homozygosity for *COMT: rs4818* (2.7% vs. 10.3%, p<0.001) and *OPRM1: rs1799971* (2.7% vs. 7.1%, p<0.001) variants. The frequencies of homozygosity for *COMT: rs4680, COMT: rs4633*, and *DRD2/ANKK1: rs1800497* variants were not significantly different between high-risk infants and Ensembl. The most common CYP2D6 and CYP2B6 metabolizer phenotypes in our patient sample were intermediate (CYP2B6: 39, 35.5%, CYP2D6: 22, 20.0%) and normal (CYP2B6: 59, 53.9%, CYP2D6: 78, 70.9%).

**Table 2:**
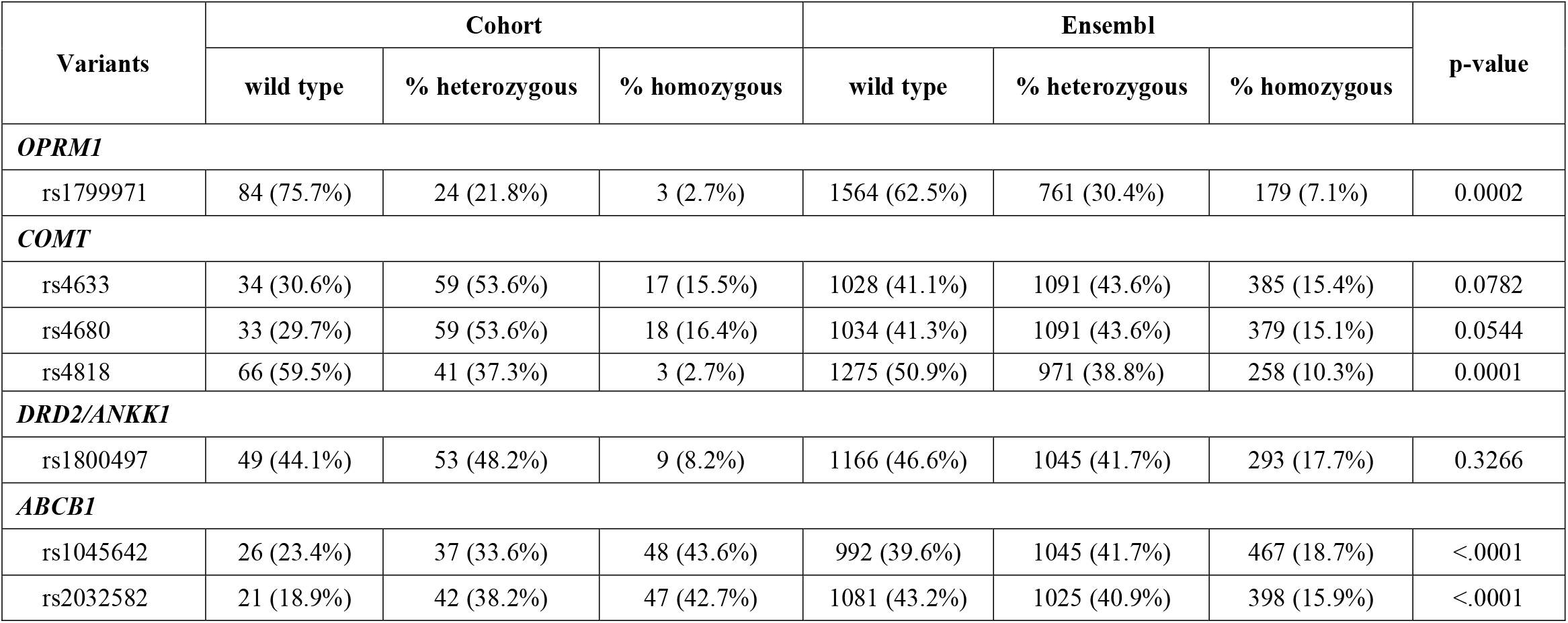
Genotype frequencies of pharmacogenomic (PGx) variants associated with opioid metabolism and altered reward/pain compared to Ensembl.

| Variants | Cohort |  |  | Ensembl |  |  | p-value |
| --- | --- | --- | --- | --- | --- | --- | --- |
|  | wild type | % heterozygous | % homozygous | wild type | % heterozygous | % homozygous |  |
| OPRM1 |  |  |  |  |  |  |  |
| rs1799971 | 84 (75.7%) | 24 (21.8%) | 3 (2.7%) | 1564 (62.5%) | 761 (30.4%) | 179 (7.1%) | 0.0002 |
| COMT |  |  |  |  |  |  |  |
| rs4633 | 34 (30.6%) | 59 (53.6%) | 17 (15.5%) | 1028 (41.1%) | 1091 (43.6%) | 385 (15.4%) | 0.0782 |
| rs4680 | 33 (29.7%) | 59 (53.6%) | 18 (16.4%) | 1034 (41.3%) | 1091 (43.6%) | 379 (15.1%) | 0.0544 |
| rs4818 | 66 (59.5%) | 41 (37.3%) | 3 (2.7%) | 1275 (50.9%) | 971 (38.8%) | 258 (10.3%) | 0.0001 |
| DRD2/ANKK1 |  |  |  |  |  |  |  |
| rs1800497 | 49 (44.1%) | 53 (48.2%) | 9 (8.2%) | 1166 (46.6%) | 1045 (41.7%) | 293 (17.7%) | 0.3266 |
| ABCB1 |  |  |  |  |  |  |  |
| rs1045642 | 26 (23.4%) | 37 (33.6%) | 48 (43.6%) | 992 (39.6%) | 1045 (41.7%) | 467 (18.7%) | <.0001 |
| rs2032582 | 21 (18.9%) | 42 (38.2%) | 47 (42.7%) | 1081 (43.2%) | 1025 (40.9%) | 398 (15.9%) | <.0001 |

**Table 3:** Genotype frequencies of variants of metabolizer phenotypes in CYP enzymes within our analytic sample.

| **Metabolizer Phenotypes** | **Analytic sample (n=111)** |  |
| --- | --- | --- |
|  | **CYP2B6** | **CYP2D6** |
| **Poor** | 8 (7.3%) | 5 (4.5%) |
| **Intermediate** | 39 (35.5%) | 22 (20.0%) |
| **Normal** | 59 (53.6%) | 78 (70.9%) |
| **Rapid** | 2 (1.8%) | n/a |
| **Ultrarapid** | 0 (0%) | 0 (0%) |
| **Indeterminate** | 2 (1.8%) | 5 (4.5%) |

PGx variants were identified in all high-risk categories and notably all patients with surgical NEC (n = 5) were homozygous for at least one variant. High-risk infants undergoing CHD surgery (OR 1.27, CI 0.47-3.46, p = 0.813, Table 4) or non-NEC abdominal surgery (OR 3.65, CI 0.79-16.85, p=0.095), or with a diagnosis of medical NEC (OR 4.66, CI 0.58-37.15, p=0.187), surgical NEC (OR 2.47, CI 0.13-47.0, p=0.581), or very low birthweight (OR 1.19, CI 0.24-5.87, p=0.999) had an increased likelihood of expressing any PGx variant, although not statistically significant.

## Discussion

In this pilot study, we examined the prevalence of PGx variants previously reported in the literature to influence opioid response among a diverse cohort of high-risk infants hospitalized at a quaternary care center. Our findings reveal that a majority of these infants were homozygous for at least one PGx variant associated with opioid metabolism and/or pain response, with notably high frequencies of *ABCB1* variants (rs1045642 and rs2032582) compared to reference populations in Ensembl. These results suggest that PGx variants may be common among critically ill infants, which may contribute to the variability in opioid response and the risk of adverse outcomes in this population. Notably, PGx variants were present across all high-risk infant categories. In adult populations, clinician awareness of PGx variant status may help optimize opioid stewardship and improve health outcomes [29,30]. Taken collectively, these findings highlight potential for personalized medicine approaches to pain management for critically ill hospitalized infants [31].

We observed a high prevalence of *ABCB1* variants (*rs1045642* and *rs2032582*) among our cohort compared to Ensembl. These variants are suggested to influence the pharmacokinetics of opioids by affecting drug transport across the blood-brain barrier [23]. Previous studies have highlighted the role of *ABCB1* polymorphisms in altering opioid efficacy and risk of adverse effects, particularly in adult populations [3,13]. Additionally, one study indicated a potential contribution of *ABCB1* variants in the development of opioid addiction, however this was demonstrated in adults of European descent [24]. Future research should focus on elucidating the functional consequences of *ABCB1* variants in high-risk infants and their potential role in guiding opioid dosing strategies.

We found a relatively low frequency of homozygosity for the *COMT: rs4818* and *OPRM1: rs1799971* variants in our cohort compared to the Ensembl database. Catechol-O-methyltransferase (COMT) is an enzyme encoded by the *COMT* gene that plays a crucial role in the methylation and degradation of catecholamines, including adrenaline, noradrenaline, and dopamine [32] As a key modulator of catecholamine levels within the pain perception pathway, COMT is instrumental in regulating pain sensitivity and has been studied for its impact on opioid response [2,3,12,21,33,34]. Variants in *COMT*, particularly *rs4680*, have been associated with differences in pain sensitivity and opioid efficacy, though the specific impact of *rs4818* remains unclear, with studies reporting mixed results [2,35]. The *OPRM1* gene encodes the mu-opioid receptor, the primary target for opioid drugs, and the *rs1799971* variant has been linked to increased risk of opioid use disorder and alterations in pain response [3,25,36–39]. However, research on the prevalence and impact of these variants in infants is limited, and our findings suggest that these variants may be infrequent in critically ill infants compared to other populations. Further research is needed to clarify the role of these variants in pediatric populations and to determine whether they should be considered in precision medicine approaches for opioid prescribing in infants.

Notably, we found that all infants with surgical NEC were homozygous for at least one PGx variant associated with opioid response. This is particularly notable given the high morbidity and mortality associated with NEC and the critical role of effective postoperative pain management in these patients [7,40]. To our knowledge, this is the first study to identify a high prevalence of opioid-related PGx variants in a group of infants with surgical NEC. These results may suggest that infants with surgical NEC may represent a high-risk group for opioid-related adverse outcomes and could benefit from targeted pharmacogenomic screening to optimize pain management strategies. Further research with larger cohorts of infants with surgical NEC are needed to confirm this finding.

In this pilot study, most infants had normal CYP2D6 and CYP2B6 metabolizer phenotypes. Additionally, we had a low prevalence of poor metabolizer phenotypes and we did not capture any infants with ultrarapid metabolizer phenotypes, and very few with poor metabolizer. However, this observed low prevalence in ultrarapid and poor metabolizer phenotypes may just be a function of our small cohort size. Further larger studies are needed to delineate the true prevalence of outlier metabolizer phenotypes among the critically ill infant population. According to the Pharmacogenomics Knowledge Base (PharmGKB), a publicly available online resource that provides comprehensive information on the effects of genetic variants, clinical guidelines, and some allele frequencies, ∼5% of Americans should have an ultrarapid metabolizer phenotype for CYP2D6, & ∼0.01-1% for CYP2B6 [41]. This may be a significant difference between infants included in our study and the overall American population, but larger cohorts are required to further investigate and confirm this difference if it exists.

Phenotypes, rather than individual variants, are used to represent predicted enzyme activity for genes like *CYP2D6* and *CYP2B6* in this study. These genes involve complex genetic variation, with multiple variants, often in the form of diplotypes (two genetic variants), contributing to the assignment of star alleles. Furthermore, *CYP2D6* is also prone to copy number variation, which can include gene duplications, deletions, and complex rearrangements [42]. To enhance clarity, we report predicted metabolizer phenotypes to summarize enzyme activity. This approach is particularly important for CYP2D6, where pharmacogenetic testing is widely implemented in clinical practice. Such testing allows clinicians to identify individuals with variant metabolizer phenotypes, enabling more informed decisions, especially regarding the prescription of codeine and tramadol, which are significantly influenced by CYP2D6 polymorphisms [43]. However, due to safety concerns, most clinicians have largely stopped prescribing codeine and tramadol altogether, reducing the need for widespread screening of these variants [44]. In this pilot study, we observed a low incidence of *CYP2D6* polymorphisms. Since neither codeine nor tramadol is typically administered in infants, these variants likely do not play a significant role in our patient population, however it is important for the child and adult population. Ultimately, this pilot study lays the groundwork for future research on pharmacogenomic variants in critically ill infants, enabling future studies to recruit larger cohorts for well-powered investigations.

CYP2B6 plays a role in the metabolism of methadone, which is commonly used for the treatment of opioid use disorder, opioid withdrawal, neonatal abstinence syndrome, and acute and chronic pain. Higher plasma S-methadone concentrations have been associated with CYP2B6 poor metabolizer statuses [26]. The Clinical Pharmacogenetics Implementation Consortium (CPIC) recently published evidence-based guidelines regarding the use of CYP2B6 testing, and due to insufficient evidence, no genotype-guided guidelines were established [45].

There are several limitations to this pilot study that should be acknowledged. First, the retrospective nature of the study design may introduce selection bias, as only infants who underwent genomic sequencing as part of their routine clinical care were included. Second, our cohort size may limit both the statistical power to detect significant associations between specific PGx variants and limit the generalizability of our findings. However, our findings underscore the need for larger studies to provide more reliable estimates of PGx variant prevalence in critically ill infant populations. Our results enable other research groups to generate estimates of prevalence and variability to sufficiently power future studies. Third, our analysis was restricted to exonic variants and previous studies highlight the potential role of intronic variants in pharmacogenomics, and their exclusion could limit our understanding of the full genetic landscape affecting opioid response [2,3]. However, the exonic variants we examined are among the most frequently reported for opioid response, supporting existing literature and adding to current knowledge. Furthermore, we recognize that the Ensembl genomic database, composed of largely European and Asian populations, does not reflect our population’s majority Hispanic/Latino ethnic makeup. Therefore, some prevalence estimates may be driven by demographic differences. Despite these limitations, our study provides preliminary prevalence estimates for opioid-related PGx variants in a high-risk infant cohort. These data highlight the need for future clinical studies examining the impact of opioid-related PGx variants across diverse pediatric populations requiring opioids in clinical care.

## Conclusion

Most high-risk infants in our pilot study were homozygous for at least one PGx variant linked to altered opioid response or reward pathways, with variant homozygosity frequencies differing from those observed in broader genetic cohorts. These findings highlight the need for larger studies that include both exonic and intronic PGx variants and use ancestrally comparable controls to assess opioid-related genetic variation in this vulnerable population. Incorporating PGx profiling into clinical practice could facilitate precision medicine approaches, enabling clinicians to tailor opioid prescribing to each patient’s unique genetic profile.

## Data Availability

All data produced in the present work are contained in the manuscript.

## Notes

### Competing Interest Statement

The authors have declared no competing interest.

### Funding Statement

This study did not receive any funding.

### Author Declarations

The study received approval from the Children's Hospital Los Angeles Review Board (CHLA-23-00335).

